# Defining the Critical Educational Components of Informed Consent for Genetic Testing: Views of US-Based Genetic Counselors and Medical Geneticists

**DOI:** 10.1101/2022.11.22.22282640

**Authors:** Miranda L. G. Hallquist, Maia J. Borensztein, Curtis R. Coughlin, Adam H. Buchanan, W. Andrew Faucett, Holly L. Peay, Maureen E. Smith, Eric P. Tricou, Wendy R. Uhlmann, Karen E. Wain, the CADRe workgroup, Kelly E. Ormond

**Affiliations:** Geisinger; Department of Genetics, Stanford University School of Medicine; Department of Pediatrics and Center for Bioethics and Humanities, University of Colorado Anschutz Medical Campus; RTI International, Genomics, Bioinformatics, and Translational Research Center; Department of Medicine, Feinberg School of Medicine, Northwestern University; Division of Genetic Medicine, Department of Internal Medicine; Department of Human Genetics; Center for Bioethics & Social Sciences in Medicine, University of Michigan; Stanford Center for Biomedical Ethics, Stanford University School of Medicine; Health Ethics and Policy Lab, Department of Health Sciences and Technology, ETH Zurich, Zurich, Switzerland

**Author notes:** Corresponding author: Miranda LG Hallquist.

## Abstract

The Clinical Genome Resource (ClinGen) <u>C</u>onsent <u>a</u>nd <u>D</u>isclosure <u>Re</u>commendation (CADRe) framework proposes that key components of informed consent for genetic testing can be covered with a targeted discussion for many conditions rather than a time-intensive traditional genetic counseling approach. We surveyed US genetics professionals (medical geneticists and genetic counselors) on their response to scenarios that proposed core informed consent concepts for clinical genetic testing developed in a prior expert consensus process. The anonymous online survey included responses to 3 (of 6 possible) different clinical scenarios that summarized the application of the core concepts. There was a binary (yes/no) question asking respondents whether they agreed the scenarios included the minimum necessary and critical educational concepts to allow an informed decision. Respondents then provided open-ended feedback on what concepts were missing or could be removed. At least one scenario was completed by 238 respondents. For all but one scenario, over 65% of respondents agreed that the identified concepts portrayed were sufficient for an informed decision; the exome scenario had the lowest agreement (58%). Qualitative analysis of the open-ended comments showed no consistently mentioned concepts to add or remove. The level of agreement with the example scenarios suggests that the minimum critical educational components for pre-test informed consent proposed in our prior work is a strong starting place for clinicians to provide targeted pre-test discussions. This may be helpful in providing consistency to the clinical practice of both genetics and non-genetics providers, and in future guideline development.

## INTRODUCTION

Historically, genetic testing was ordered by genetics specialists and pre-test genetic counseling has been a time intensive process <u>^1^</u>, likely providing more information than many patients need ^2,3^. The Clinical Genome Resource (ClinGen) <u>C</u>onsent <u>a</u>nd <u>D</u>isclosure <u>Re</u>commendation (CADRe) framework proposes that many pre-test conversations can occur with a shorter targeted discussion, utilizing the key components of informed consent ^4^. These targeted discussions have the potential to provide patients with information necessary to make a decision about testing and decrease time providers spend on consent conversations. There are examples of published criteria of concepts to cover in traditional pre-test genetic counseling ^5,6^, however there is no clear consensus on what educational content to include in a targeted consent discussion. Such a consensus would be useful for both genetics and non-genetics health care providers who conduct pre-test informed consent discussions with patients and families.

As a first step, a prior study used a modified Delphi approach with clinical genetics and bioethics experts to establish consensus on the minimum critical educational components of an informed consent process^7^. Topics identified as important in any pre-test conversation about clinical genetic testing were: (1) genetic testing is voluntary; (2) purpose of the test and what it tests for; (3) what results will be returned; (4) other potential results and options for choice (e.g., secondary findings); (5) how, if at all, prognosis and management could be impacted by the results; (6) results may be important to share with relatives and could impact family members in different ways (their health, emotions, or relationships); (7) test limitations and next steps; (8) to whom results are reported; (9) potential risk for genetic discrimination and/or stigma and protections provided by relevant federal and state laws. As expert opinions can differ from that of the general population of practicing providers, the current study aimed to assess to what degree US-based clinical genetics providers (genetic counselors (GC) and medical geneticists) agreed with the previously developed list of minimum critical educational components of informed consent.

## SUBJECTS and METHODS

### Recruitment

In February and March 2020, two recruitment emails (one invitation and one reminder) were sent to GCs and medical geneticists. GCs were emailed through the American Board of Genetic Counselors (ABGC) delegates list (N = 4,605). There is not a similar email list for medical geneticists. They were identified using a list of members of the American College of Medical Genetics and Genomics (ACMG) and searching for a publicly available email address (N = 177) and through identifying medical geneticists at major medical centers who had a publicly available email address (N=413). Respondents who self-identified as a GC or medical geneticist, had obtained informed consent for clinical genetic testing outside of being a trainee, and worked in the United States were included.

### Survey Instrumentation

The anonymous online survey was administered in Qualtrics ^8^ (Qualtrics, Provo, UT), and provided a description of the CADRe targeted discussion pre-test framework ^4^ (Supplemental Methods), gathered participant demographics, and presented example scenarios (Table 1, Supplemental Methods) that were developed by the study team. Each of the six scenarios included short case descriptions and discussion points corresponding to the consensus concepts previously identified ^7^. Participants were randomized to three scenarios, minimizing survey completion burden. The scenarios used a variety of clinical testing indications and conditions for which a targeted discussion was suggested in CADRe’s prior work ^4,9^. Each scenario included a yes or no question: “Does this scenario include the *minimum necessary and critical information* to aid a patient in making an informed decision/consent?”. Two optional open-ended questions followed: “What topic(s), if any, is critically missing?” and “What topic(s), if any, would you remove as non-critical?”. Finally, respondents were shown the complete list of consent concepts identified in the prior study^7^ to provide open-ended feedback.

**Table 1:** Example Scenarios from Survey (see supplemental methods for all scenarios)

|  |  |  |
| --- | --- | --- |
| **Core Concept for Informed Consent**<br>Critical and minimally necessary concepts identified through expert consensus process in prior study<sup>7</sup> | **Long QT Syndrome Scenario**<br>30-year-old with a clinical diagnosis of LQTS due to personal medical history and consistent family history. Discussion of genetic testing using multigene panel related to LQTS. | **Hereditary Breast and Ovarian Cancer Scenario**<br>25-year-old with no personal history of cancer. Family history significant for multiple HBOC-related cancers; family members have not pursued genetic testing. Discussion of genetic testing using multigene panel related to HBOC. |
| **Genetic testing is voluntary** | Genetic testing is optional | Genetic testing is optional |
| **Purpose of the test and what it tests for** | We are doing this test to look for a genetic cause of your LQTS. The genetic test evaluates the genes associated with LQTS. | We are doing this test to look for a genetic predisposition to cancer based on your family history of cancer. The genetic test evaluates the genes associated with hereditary breast and ovarian cancers. |
| **What results will be returned** | The test may be positive, meaning that we found a variant in a gene that is responsible for the LQTS. | The test may be positive, meaning that we found a variant in a gene that puts you at greater risk for developing breast, ovarian, and potentially other cancers, and likely explains your family history of cancer. |
| **Other potential results and options for choice (e.g., secondary findings)** | Not applicable | It is possible to cast a wider net and test for additional genes associated with hereditary cancers. But the current test will only focus on genes related to breast and ovarian cancer risks. |
| **How, if at all, prognosis and management could be impacted by the results** | While you are already being treated for LQTS, a positive test could refine your medical management if a gene specific treatment becomes known. In some cases, a specific gene diagnosis can also change what we know about your prognosis. | A positive test result for hereditary breast and ovarian cancer would come with screening recommendations depending on the gene identified, such as earlier and more frequent mammograms. For some genes, a surgery to remove the ovaries and reduce ovarian cancer risk may also be recommended after age 35. |
| **Results may be important to share with relatives and could impact family members in different ways (their health, emotions, or relationships)** | The results may impact your family in different ways (their health, emotions, or relationships), and you may want to share the results. For example, positive test results could be used for your family members to find the cause of their symptoms, to predict risk for relatives who do not have symptoms, and for family planning. | The results may impact your family in different ways (their health, emotions, or relationships), and you may want to share the results. For example, positive results on the test could be used for your family members to find the cause of their cancers, to predict risk for relatives who do not have symptoms, and for family planning. |
| **Test limitations and next steps** | The test has limits. Even though you have a clinical diagnosis of LQTS, the test may be negative, meaning that we may not find a genetic cause of your LQTS in the genes tested. Lastly it may be inconclusive, meaning that a variant was found but we do not know if it causes your LQTS or is normal genetic variation. We would discuss any possible next steps after results are available. | The test has limits. We may not find a genetic change associated with cancer risk in the genes tested, a 'negative' result'. It would still be possible that there is a genetic predisposition to cancer in your family members that you did not inherit, and they could consider testing. It would also be possible that there is a genetic cause for the cancers in your family that we can't test for, and you would still be at an increased risk for cancer. If we do not find a genetic change in you, we will still recommend cancer screening based on your family history. Lastly it may be inconclusive, meaning that a variant was found but we do not know if it increases your risk for cancer or is normal genetic variation. In either case we would discuss any possible next steps after results are available. |
| **To whom results are reported** | The results would be reported directly to me and I will contact you with them by [manner and approximate timeline]. | The results would be reported directly to me and I will contact you with them by [manner and approximate timeline]. |
| **Potential risk for genetic discrimination and/or stigma and protections provided by relevant federal and state laws** | While genetic information is protected against discrimination of employment or health insurance by federal law, it is not protected against discrimination for long term care or disability, or life insurance. This is most relevant for relatives without symptoms who choose to have genetic testing | While genetic information is protected against discrimination of employment or health insurance by federal law, it is not protected against discrimination for long term care or disability, or life insurance. This is most relevant for people without symptoms who choose to have genetic testing. |

Abbreviations: LQTS = Long QT Syndrome HBOC = Hereditary Breast and Ovarian Cancer

### Data Analysis

We used SPSS version 26 ^10^ (2019) to conduct a descriptive analysis of frequency and means. For each consent scenario, we calculated the frequency of agreement for the total group and each professional group. Agreement was calculated as the proportion of respondents who agreed that the scenario included the minimum necessary and critical information for an informed decision. Chi-square tests assessed statistical differences in scenario agreement between GCs and medical geneticists and between clinicians with and without clinical experience in the medical specialty for a given scenario (e.g., cardiogenetics provider and Long QT Syndrome (LQTS) scenario). A significant result was defined as p ≤ 0.05.

Open-ended comments were reviewed by three investigators (MLGH, MJB, KEO), and a code book was inductively developed from the data. Codes were applied by two coders until consensus was met (MLGH, KEO). Codes were evaluated for each scenario separately, as well as for themes that emerged across all scenarios.

## RESULTS

A total of 238 respondents are included; 215 completed all scenarios and demographic questions, and 23 completed at least one scenario. Response rate was 9% for medical geneticists (54/590), and 4% for GCs (184/4605). Table 2 summarizes the demographics of respondents, who were primarily GCs (77%), identified as female (86%) and had <10 years of experience (54%). Informed consent for genetic tests was regularly obtained by 78% of respondents, with 42% indicating frequency of 21 or more patients per month. The majority of respondents reported having current or past experience working in general/medical genetics (67%) or cancer genetics (50%), with many respondents also having experience in reproductive genetics (44%), neurogenetics (23%), and cardiovascular genetics (20%).

**Table 2:** Survey Demographics.

|  |  | **Total**<br>**Respondents**<br>**(n=215\*)** | **Genetic**<br>**Counselors**<br>**(n=163)** | **Medical**<br>**Geneticists**<br>**(n=52)** |
| --- | --- | --- | --- | --- |
| **Gender** | Female | 184 (85.6%) | 156 (95.7%) | 28 (54%) |
|  | Male | 29 (13.5%) | 6 (4%) | 23 (44%) |
|  | Prefer not to say | 2 (1%) | 1 (0.6%) | 1 (2%) |
| **Age (years)** | ≤24 | 6 (3%) | 6 (4%) | - |
|  | 25-29 | 47 (22%) | 47 (29%) | - |
|  | 30-34 | 41 (19%) | 39 (24%) | 2 (4%) |
|  | 35-39 | 27 (13%) | 18 (11%) | 9 (17%) |
|  | 40-44 | 21 (10%) | 17 (10%) | 4 (8%) |
|  | 45-49 | 21 (10%) | 12 (7%) | 9 (17%) |
|  | 50-54 | 16 (7%) | 12 (7%) | 4 (8%) |
|  | 55-59 | 13 (6%) | 5 (3%) | 8 (15%) |
|  | 60-64 | 10 (5%) | 4 (3%) | 6 (12%) |
|  | 65-69 | 9 (4%) | 3 (2%) | 6 (12%) |
|  | 70+ | 4 (2%) | - | 4 (8%) |
| **Years of experience** | <1 | 16 (7%) | 15 (9%) | 1 (2%) |
|  | 1-4 | 53 (25%) | 47 (30%) | 6 (12%) |
|  | 5-9 | 47 (22%) | 38 (23%) | 9 (17%) |
|  | 10-14 | 27 (13%) | 17 (10%) | 10 (19%) |
|  | 15-19 | 20 (9%) | 17 (10%) | 3 (6%) |
|  | 20-25 | 19 (9%) | 17 (10%) | 2 (4%) |
|  | 25+ | 33 (15%) | 12 (7%) | 21 (40%) |
| **Work setting\*\*** | University medical center | 113 (53%) | 75 (46%) | 38 (73%) |
|  | Public hospital/medical facility | 37 (17%) | 29 (18%) | 8 (15%) |
|  | Private hospital/medical facility | 40 (19%) | 30 (18%) | 10 (19%) |
|  | Physician's private practice | 10 (5%) | 9 (6%) | 1 (2%) |
|  | Private practice (self-employed) | 4 (2%) | 4 (3%) | - |
|  | Academic diagnostic laboratory | 4 (2%) | 3 (2%) | 1 (2%) |
|  | Commercial diagnostic laboratory | 15 (7%) | 12 (7%) | 3 (6%) |
|  | Research development/biotechnology company | 3 (1%) | 3 (2%) | - |
|  | Government organization/agency | 11 (5%) | 9 (6%) | 2 (4%) |
|  | Internet/website company | 2 (1%) | 2 (1%) | - |
| **Direct patient care** | Yes | 189 (88%) | 140 (86%) | 49 (94%) |
| **Work with patients** | Regularly see patients and obtain consent for genetic testing | 167 (78%) | 124 (76%) | 43 (85%) |
|  | Occasionally see patients and obtain consent for genetic testing | 23 (11%) | 16 (10%) | 7 (14%) |
|  | Do not currently, but have previously seen patients and obtained consent for genetic testing | 24 (11%) | 23 (14%) | 1 (2%) |
| **Frequency of consents (per month)** | 0 | 24 (11%) | 23 (14.%) | 1 (2%) |
|  | <10 | 52 (24%) | 30 (18%) | 22 (42%) |
|  | 11-20 | 49 (23%) | 38 (23%) | 11 (21%) |
|  | 21-30 | 39 (18%) | 29 (18%) | 10 (19%) |
|  | >30 | 51 (24%) | 43 (26%) | 8 (15%) |
| **Specialty area\*\***<br>Currently practicing<br>[Ever practiced^] | **Reproductive genetics** | 49 (23%)<br>[95 (44%)] | 41 (25%)<br>[76 (47%)] | 8 (15%)<br>[19 (37%)] |
|  | **General / Medical genetics** | 103 (48%)<br>[144 (67%)] | 60 (37%)<br>[95 (58%)] | 43 (83%)<br>[49 (94%)] |
|  | **Cardiovascular genetics** | 36 (17%)<br>[44 (21%)] | 19 (12%)<br>[23 (14%)] | 17 (33%)<br>[21 (40%)] |
|  | **Cancer genetics** | 82 (38%)<br>[108 (50%)] | 65 (40%)<br>[88 (54%)] | 17 (32%)<br>[20 (39%)] |
|  | **Neurogenetics** | 42 (20%)<br>[51 (24%)] | 24 (15%)<br>[30 (18%)] | 18 (35%)<br>[21 (40%)] |
|  | **Research** | 45 (21%)<br>[65 (30%)] | 26 (16%)<br>[40 (25%)] | 19 (37%)<br>[25 (48%)] |
|  | **Predictive Genomic Medicine** | 25 (12%)<br>[34 (16%)] | 13 (8%)<br>[21 (13%)] | 12 (23%)<br>[13 (25%)] |
|  | **Laboratory** | 27 (13%)<br>[41 (19%)] | 18 (11%)<br>[29 (18%)] | 9 (17%)<br>[12 (23%)] |
|  | **Other medical specialties** | 22 (10%)<br>[35 (16%)] | 15 (9%)<br>[24 (15%)] | 7 (14%)<br>[11 (21%)] |
|  | **Non-patient facing role only** | 16 (7%)<br>[23 (10.7%)] | 16 (10%)<br>[23 (14.1%)] | - |

\*Respondents included in this table completed the demographics questions at the end of the survey. This is a subset of the total 238 respondents (184 GCs and 54 medical geneticists) included in analysis of scenarios based on having completed at least one scenario.
\*\*Total more than 100% because respondents could select multiple options
^ Ever practiced includes respondents who reported currently practicing or having previous experience of over a year in a given specialty.

As shown in Table 3, 58%-74% of respondents in each scenario agreed that the identified concepts portrayed were sufficient for an informed decision. There were no statistically significant differences between the agreement of GCs and medical geneticists for any scenario. Concordance between participants’ specialty and the content of the scenario only led to statistically significant differences in agreement for the LQTS scenario. Providers with cardiovascular genetics experience were significantly less likely to agree that the concepts in the scenario were sufficient for informed consent than those without cardiogenetics experience (48% vs. 78%, Pearson chi-square (df 1, N=110) = 8.99, p = .003).

**Table 3:**
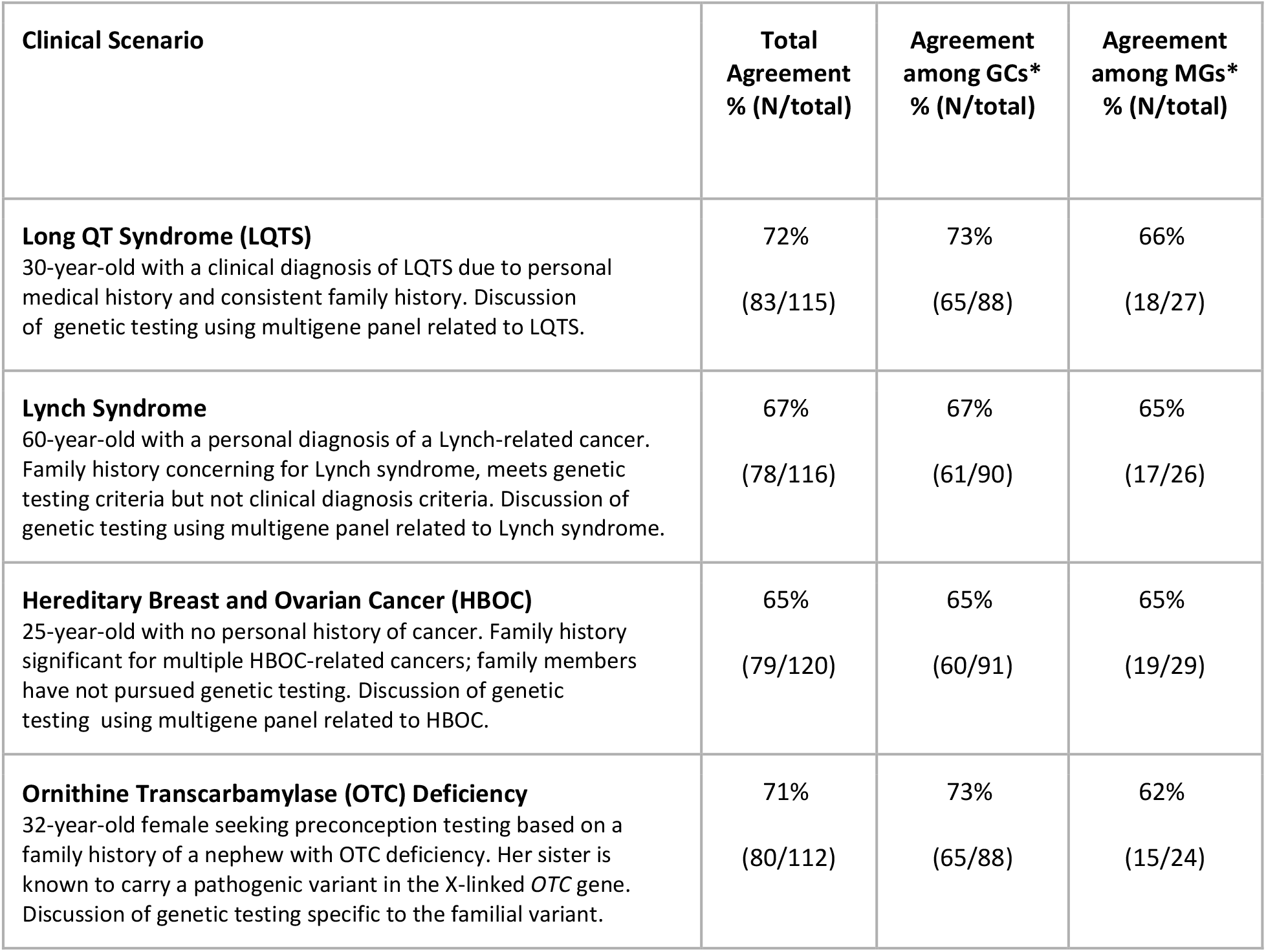
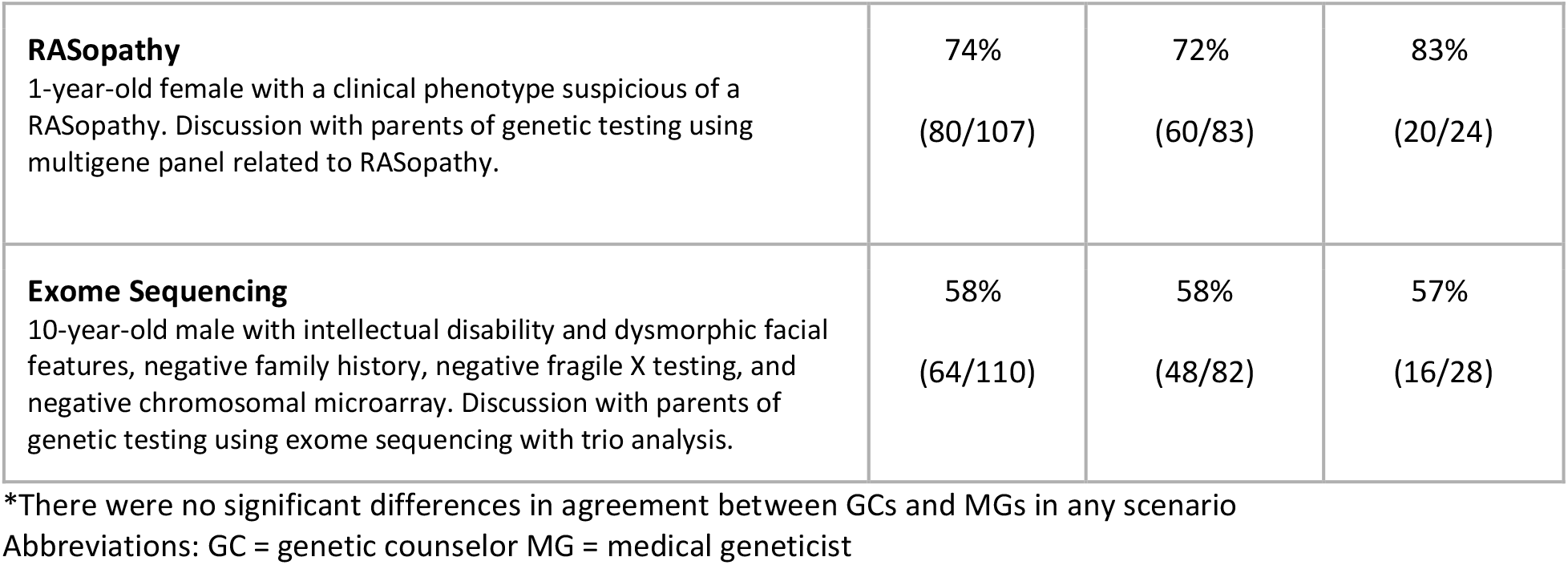
Agreement that concepts portrayed in scenario were sufficient for an informed decision.

Analysis of the open-ended comments (Table 4) provided insight into the respondents’ thought process; 186 respondents (78%) commented at least once, with each scenario receiving 39-59 comments on missing concepts and 13-29 comments on removable concepts. Respondents to the exome scenario highlighted the topic of unanticipated findings, including both secondary or incidental findings such as misattributed parentage through trio analysis, and reminders that an exome test “does not look at all the genes”. Unique to the hereditary breast and ovarian cancer scenario, where an unaffected individual is pursuing testing based on a family history of cancer, one of the most common missing topics was that the test results would be more informative in a relative with a personal history of cancer. Particularly common in the LQTS example, where testing was performed on an individual who already had a clinical diagnosis, was the concept that negative genetic testing does not change clinical management. Finally, each of the scenarios had some responses (5-15) commenting on a desire for more information about general genetics knowledge (e.g., inheritance), cost and logistics, medical management, family implications, and residual risks after negative testing.

**Table 4:** Qualitative comments on suggested additional concepts for an informed decision.

| Clinical Scenario<br>(n commented/n total respondents) | Four most common themes for missing concepts within each scenario (most to least frequent) | Example Qualitative Comments (participant specialty concordance and role) |
| --- | --- | --- |
| **Long QT Syndrome (LQTS)**<br>(39/115)<br>30-year-old with a clinical diagnosis of LQTS due to personal medical history and consistent family history. Discussion of genetic testing using multigene panel related to LQTS. | Residual risk if negative<br>• Risk of phenotype<br>• Risk of genetic cause not identified by testing | “A negative test result does not rule out a genetic cause.” (Non-cardio GC)<br>“A negative test result does not rule out his diagnosis of LQTS.” (Cardio GC) |
|  | Knowledge of genetics and genetic testing<br>• Basic genetics (e.g., inheritance)<br>• Changes in knowledge over time (e.g., gene discovery, variant interpretation, testing technology) | “Additional genetic testing options may be available in the future.” (Non-cardio GC)<br>“Brief overview of genetics, so the patient knows what a gene is.” (Non-cardio GC) |
|  | Family implications<br>• Family member risk for phenotype (if proband negative)<br>• Family member risk for genotype (if proband positive) | “Screening for family members would be based on the family history if negative results.” (Cardio MG)<br>“The main benefit of genetic testing in this scenario would be for unaffected or unscreened family members to know [the] familial mutation to pursue risk evaluation or for PGD.” (Cardio GC) |
|  | Cost and logistics<br>• Out of pocket cost<br>• Insurance coverage and logistics (e.g., prior authorization)<br>• Sample logistics | “Insurance coverage and any possible out of pocket costs involved with testing.” (Non-Cardio GC)<br>“How long will it take to complete the testing.” (Cardio MG) |
| **Lynch Syndrome**<br>(59/116)<br>60-year-old with a personal diagnosis of a Lynch-related cancer. Family history concerning for Lynch syndrome, | Knowledge of genetics and genetic testing<br>• Basic genetics (e.g., inheritance)<br>• Changes in knowledge over time (e.g., gene discovery, variant interpretation, testing technology) | “Basic ‘Genetics 101’ - DNA, genes, chromosomes.” (Non-cancer GC)<br>“Updated interpretation over time (upgrade or downgrade if VUS).” (Non-cancer GC) |
| meets genetic testing criteria but not clinical diagnosis criteria. Discussion of genetic testing using multigene panel related to Lynch syndrome. | Residual risk if negative<br>• Risk of phenotype<br>• Risk of genetic cause not identified by testing | “Screening for the various Lynch syndrome cancers should be considered/performed if there is a negative result.” (Cancer MG)<br>“If testing doesn't find anything, risk is still elevated over population risk.” (Non-cancer GC) |
|  | Cost and logistics<br>• Out of pocket cost<br>• Insurance coverage and logistics (e.g., prior authorization)<br>• Sample logistics | “How testing will be performed (i.e. sample type and associated risks).” (Non-cancer GC)<br>“Cost of genetic testing/insurance coverage.” (Cancer GC) |
|  | Management implications<br>• Changes in management if positive (if any)<br>• Changes in management if negative (if any) | “Implications for change in cancer therapy if positive (possible immunotherapy clinical trial).” (Cancer GC)<br>“More detail about frequency [of screening] and what surgeries would be important.” (Cancer GC) |
| **Hereditary Breast and Ovarian Cancer (HBOC) (53/120)**<br>25-year-old with no personal history of cancer. Family history significant for multiple HBOC-related cancers; family members have not pursued genetic testing. Discussion of genetic testing using multigene panel related to HBOC. | Genetic testing approach in family | “A statement about testing an affected family member (and ideally first) as the best approach is needed so that the patient's test results could be more fully interpreted.” (Cancer MG)<br>“An affected family member would be the best person to start familial testing.” (Non-cancer GC) |
|  | Knowledge of genetics and genetic testing<br>• Basic genetics (e.g., inheritance)<br>• Changes in knowledge over time (e.g., gene discovery, variant interpretation, testing technology) | “There are 3 main causes of cancer- sporadic, environmental and hereditary. Most are not hereditary.” (Cancer GC)<br>“New genes may be discovered later that are not on the current panel.” (Non-cancer MG) |
|  | Chance of positive result that did not have clear management guidelines | “Not all genes are well understood and consensus guidelines may not be available to guide screening / management.” (Cancer GC)<br>“I feel that this information does not make clear the possibility of identifying a significant risk for a cancer *other than* the ones the patient described in the family history, which |
|  |  | could in turn lead to entirely unexpected recommendations for screening & preventive measures (a classic example is CDH1).” (Cancer GC) |
|  | Genetic testing choices related to panel size | “I also would offer the broadest panel available to the patient, unless they specifically wanted a panel limited to certain types of cancer only.” (Non-cancer GC)<br>“Exactly what genes or conditions are being tested (BRCA1/2 only vs panel).” (Cancer GC) |
| **Ornithine Transcarbamylase (OTC) Deficiency (44/112)**<br>32-year-old female seeking preconception testing based on a family history of a nephew with OTC deficiency. Her sister is known to carry a pathogenic variant in the X-linked *OTC* gene. Discussion of genetic testing specific to the familial variant. | Knowledge of genetics and genetic testing<br>• Basic genetics (e.g., inheritance)<br>• Changes in knowledge over time (e.g., gene discovery, variant interpretation, testing technology) | “There should be a discussion of mode of inheritance and prior risk of being a carrier.” (MG)<br>“Other genetic risk may still be inherited”. (MG) |
|  | Management implications<br>• Changes in management if positive (if any)<br>• Changes in management if negative (if any) | “I would discuss more about OTC deficiency, in males and females, and give her more detail about possible risks during a pregnancy if she is a carrier.” (MG)<br>“Options for prenatal diagnosis including the option to terminate an affected pregnancy.” (Peds GC) |
|  | Cost and logistics<br>• Out of pocket cost<br>• Insurance coverage and logistics (e.g., prior authorization)<br>• Sample logistics | “Information on what testing involves (e.g. blood draw, saliva sample).” (Peds GC)<br>“Ideally there would be mention of insurance coverage and any possible out of pocket costs involved with testing.” (Peds GC) |
|  | Reproductive considerations | “Also would have talked in more detail about available technologies that could be used to test the pregnancy or to do PGT-M” (Peds GC)<br>“The types of tests that would be offered to test a conception/pregnancy in the event she was positive for the familial variant: PGT with IVF, CVS, amniocentesis.” (Non-peds GC) |
| **RASopathy (37/107)**<br>1-year-old female with a clinical phenotype suspicious of a RASopathy. Discussion with parents of genetic testing using multigene panel related to RASopathy. | Knowledge of genetics and genetic testing<br>Basic genetics (e.g., inheritance)<br>Changes in knowledge over time (e.g., gene discovery, variant interpretation, testing technology) | “Variant interpretation represents our best understanding of pathology at this time. Some variants can be reclassified in the future.” (MG)<br>“Explaining [autosomal dominant inheritance] briefly, that her features could be de novo or inherited/possibility that a parent also has the same thing.” (Non-peds GC) |
|  | Cost and logistics<br>Out of pocket cost<br>Insurance coverage and logistics (e.g., prior authorization)<br>Sample logistics | “Discussion of cost of testing/insurance coverage, as this may impact willingness to do the testing.” (Non-peds GC)<br>“Discuss potential costs should insurance not cover the expense of testing.” (Peds GC) |
|  | Family implications<br>Family member risk for phenotype (if proband negative)<br>Family member risk for genotype (if proband positive) | “Possible need for parental testing to determine significance of VUS or recurrence risk.” (Non-peds GC)<br>“Implications for family members, including discussion of inheritance.” (Peds GC) |
|  | Residual risk if negative<br>Risk of phenotype<br>Risk of genetic cause not identified by testing | “A negative result does not rule out a genetic basis for their child's disease.” (Non-peds GC)<br>“This testing could miss some conditions that were not tested for.” (MG) |
| **Exome Sequencing\* (55/110)**<br>10-year-old male with intellectual disability and dysmorphic facial features, negative family history, negative fragile X testing, and negative microarray. Discussion with parents of genetic testing using exome sequencing with trio analysis. | Knowledge of genetics and genetic testing<br>Basic genetics (e.g., inheritance)<br>Changes in knowledge over time (e.g., gene discovery, variant interpretation, testing technology) | “Options for future analysis, including the fact that a negative or inconclusive result could be different as our understanding of genes and diseases advances.” (Peds GC)<br>“Exome sequencing cannot detect all, in fact many, genetic mutations that could lead to disease in genes analyzed and incidental mutations in genes not analyzed would not be found.” (Peds GC) |
|  | Unanticipated results<br>Unanticipated secondary findings<br>Unanticipated family relationships due to trio/duo testing | “May reveal another condition unrelated to the reason for which he is being tested.” (Peds GC)<br>“Unexpected family relationships needs to be further defined. For example, we could |
|  |  | learn that one of you are not the patient's biologic parent." (MG) |
|  | Cost and logistics<br>• Out of pocket cost<br>• Insurance coverage and logistics (e.g., prior authorization)<br>• Sample logistics | "Cost is a huge issue, that should be part of the consent process." (Peds GC)<br>"I'd add the procedure required (i.e. venipuncture)." (Peds GC) |
|  | Management implications<br>• Changes in management if positive (if any)<br>• Changes in management if negative (if any) | "Negative results would not change the services he is already receiving." (Peds GC)<br>"A positive test could, in some cases, not provide any additional information about services or health concerns." (Peds GC) |
|  | Residual risk if negative<br>• Risk of phenotype<br>• Risk of genetic cause not identified by testing | "The test is not perfect. It may miss the cause of the patient's disease or the genetic cause of his illness has not yet been discovered." (MG)<br>"Negative results do not rule out the possibility of a genetic condition." (Non-peds GC) |

\*Five themes are noted for the exome sequencing scenario as management implications and residual risk if negative had equal frequency, both being fourth most common.
Abbreviations: GC = genetic counselor MG = medical geneticist

Of the 126 (53%) respondents who provided feedback regarding the full list of informed consent concepts, 22 indicated that the list was adequate. Additions most commonly suggested were: cost (16), changing understanding of genetics over time (9), management (8), and test limitations (7). The only concept commonly suggested for removal was the Genetic Information Nondiscrimination Act (GINA) (16), as this was seen as only relevant in some testing indications. All other concepts were mentioned five or fewer times each.

## DISCUSSION

This is one of the first studies to examine how practicing genetics professionals responded to a proposal on the core content for targeted pre-test discussions. For five of the six scenarios, at least 65% of survey respondents agreed that each example clinical scenario met the threshold of *minimal necessary and critical* components. There was somewhat lower agreement among respondents to the exome scenario, with 58% agreement that the scenario met the minimal necessary and critical criteria. Our use of a binary yes/no question to assess agreement with the critical concept list may underestimate the proportion of respondents who agreed generally with the concepts but had minor suggestions. Some respondents suggested word or phrasing changes, making it difficult to assess if lack of agreement was related to the scenario phrasing or the underlying concepts. Furthermore, some suggestions were additions of topics that we had considered to be already conceptually included. For example, we included the concept that the results of the genetic testing would have implications for management and some respondents suggested that specific management options should be reviewed in detail.

When presented with the complete list of the consent concepts identified in the prior study^7^, no concepts were consistently suggested for addition or removal. Rather, different concepts were supported by a minority of respondents, suggesting that there were no additional topics that were universally suggested for inclusion in informed consent. Together, the level of agreement and lack of consistent additional concepts suggests that the minimum critical educational components for pre-test informed consent proposed in our prior work is a strong starting place for clinicians who provide targeted informed consent discussions.

The qualitative comments for individual scenarios suggest that critical topics and level of discussion depth vary slightly based on the clinical indication. For example, if someone is unaffected with a relevant disease, some respondents felt it may be important to emphasize the limitations of a negative result; whereas, for someone who has a clinical diagnosis of a condition, it may be important to detail that negative results do not change their diagnosis or management. The latter point seems particularly prominent in conditions in which management occurs for individuals with a clinical diagnosis regardless of genetic status, such as LQTS. More cardiology providers commented that a negative result would not change the clinical diagnosis or alter the recommended LQTS screening and management than did non-cardiology providers, potentially explaining the emphasis that cardiology providers place on this component of informed consent compared to providers in other specialties. While the additional concepts suggested for the exome sequencing scenario were broadly included in the minimal necessary list, such as options related to secondary findings, the lower agreement with this scenario suggests that exome sequencing may have additional complexities that warrant inclusion in a targeted discussion or in supportive education materials.

Regarding concepts that might be removed from the core concept list, respondents noted that a discussion of the potential for genetic discrimination was not always needed, especially in the Lynch syndrome diagnostic testing scenario. Given that this concept was also one that did not have clear consensus in the prior study ^7^, it reinforces that a discussion about potential genetic discrimination and legal protections is most relevant for predictive testing scenarios ^11^.

Across all scenarios, respondents mentioned that topics related to providing basic information about genetics were missing from the concept list. While traditional genetic counseling has historically included a review of genes, chromosomes, inheritance patterns, and the specifics of the testing technology, recent literature and our own prior study suggest that this may be more information than many patients need, especially during an informed consent process ^2,3,7^. Since the value of providing basic genetics educational information has not been supported by empirical research, these findings may represent provider views of traditional norms. Furthermore, some respondents highlighted the importance of explaining how our knowledge of genetics changes over time, suggesting that this may be a more useful way to frame some genetic testing conversations.

A brief consensus list of concepts critical for informed consent before genetic testing, like the one we have tested in this study, could guide targeted pre-test discussions. By standardizing and shortening the core information that all patients receive, providers may be better able to leave space to further tailor consent conversations to patients’ specific questions and concerns. As such, this list could support current service delivery models aimed at streamlining the consent process. For example, such a list may be useful for specialists ordering genetic testing prior to referring to genetic counseling, which is becoming increasingly common as genetic services expand around the globe^12^. While some specialties, such as oncology, have developed guidelines for genetic testing consent^5^, these core concepts will be especially useful where there are no available guidelines or providers are newer to genetic testing. Furthermore, a list of minimally necessary concepts may aid in the development of pre-test educational materials such as videos or chatbots.

As proposed in the CADRe framework^4^, a model that supports targeted pre-test discussion allows for post-test discussions to be guided by the results of genetic testing, with the opportunity to address details about risks and management that are most relevant to the patient. This, in turn, may support genetics providers dedicating time to helping patients navigate positive or uncertain results, where their expertise may be most valuable. Future work to understand how other stakeholders, especially diverse patient populations, non-genetics providers, and genetics experts internationally, respond to our proposed consensus list for minimally necessary concepts will be imperative to successfully implement pre-test informed consent in a targeted manner.

### Limitations

While we had 238 respondents, the low response rate (<10%) limits the generalizability of our results. It is possible that if a broader group of individuals participated, we would have seen different levels of agreement with the proposed list. The low response rate also precluded us from completing additional sub-analyses to see if, for example, years in the field or number of consents per month was associated with scenario agreement. The majority of respondents were genetic counselors, though we identified no significant differences between genetic counselors and medical geneticists. Finally, this study only surveyed clinical genetics providers in the US.

### Conclusion

Respondents to the survey generally supported the expert consensus list developed in our prior work, ^7^ suggesting that it is a reasonable starting point when outlining the educational concepts to be included in a targeted discussion for pre-test informed consent. This study also reinforces that different clinical situations and testing will require additional tailored information in many cases. Importantly, the process of informed consent includes a bidirectional conversation between clinician and patient. Having a list of key educational concepts as a starting place for discussion does not imply that consent conversations should only address these concepts.

Providing psychosocial support, addressing patient questions, and discussing test logistics are critical parts of the process. Rather, we propose that clinicians might use this list as a way to identify which of many educational topics are critical for most patients, and then follow up with specific additional information as needed based on the clinical scenario and the patient’s interests and questions, thus tailoring discussions to best match patients’ needs.

## Data Availability

The data that support the findings of this study are available within the article and/or are available on reasonable request from the corresponding author, M.L.G.H.

## ACKNOWLEDGEMENTS

Thanks to Erin Ramos and Nicole Lockhart at NHGRI, and all the remaining members of the CADRe workgroup: Laura Hercher, Howard P. Levy, Julianne M. O’Daniel, Juliann M. Savatt, Melissa Stosic, and Hannah Wand. The work described was based on input from the CADRe working group, with additional members having contributed conceptually at various stages in the development of the rubrics, including: Kyle B. Brothers, Louanne Hudgins, Seema M. Jamal, Dave Kaufman, and Myra I. Roche. Finally, we are grateful to members of the Stanford Center for Biomedical Ethics writing seminar for comments on an earlier version of this paper.

## AUTHOR CONTRIBUTIONS

Conceptualization: K.E.O, M.J.B, M.L.G.H, A.H.B, W.A.F, H.L.P, M.E.S, E.P.T, W.R.U, K.E.W., C.R.C. Data Curation: M.L.G.H, K.E.O, M.J.B.; Formal Analysis: M.L.G.H, M.J.B, K.E.O; Funding Acquisition: K.E.O, A.H.B, W.A.F; Investigation: K.E.O, M.J.B.; Methodology: K.E.O, M.J.B, M.L.G.H, A.H.B, W.A.F, H.L.P, M.E.S, E.P.T, W.R.U, K.E.W., C.R.C.; Project administration: M.J.B. Supervision: K.E.O, M.L.G.H, A.H.B, C.R.C; Visualization: K.E.O, M.J.B., M.L.G.H. Writing - original draft: M.J.B, M.L.G.H, K.E.O; Writing - review and editing: K.E.O, M.J.B, M.L.G.H, A.H.B, W.A.F, H.L.P, M.E.S, E.P.T, W.R.U, K.E.W., C.R.C.

## ETHICAL APPROVAL

The Stanford IRB reviewed this study as exempt, and all aspects adhered to the principles in the Declaration of Helsinki. All participants reviewed an informed consent document before choosing to participate in this study.

## Supplemental Methods

### Survey Instrument

#### Survey Introduction, Study Description

Despite there being much discussion about the need for a streamlined, ‘targeted discussion’ approach to consent preceding clinical genetic testing, there are no practice guidelines to define the critical elements of informed consent for these situations. The goal of this study, conducted by the ClinGen working group for Consent & Disclosure Recommendations (CADRe; clinicalgenome.org), is to identify the key elements to include in a consent conversation in order to allow patients to make an informed decision about genetic testing.

This study is the third stage of a larger Delphi consensus modeling study performed by CADRe. Prior stages presented a small group of experts in clinical genetics and bioethics with a list of >75 concepts of informed consent, and used 2 rounds of group ranking to refine the list down to a smaller proposed list of concepts critical to obtaining informed consent in clinical genetic testing.

<u>The purpose of the current study is to determine the extent to which the broader community of clinical genetics colleagues (genetic counselors and medical geneticists practicing in the US) agree with the Delphi findings.</u>

This survey should take 10-20 minutes to complete.

- In Section 1 you will be asked three short eligibility questions.
- In Section 2 you will read and answer questions about 3 short clinical scenarios. The scenarios describe minimum and critical components of a ‘targeted discussion’ approach to genetic testing consent.In Section 3 you will be shown a list of summary concepts that were included in the scenarios.
- Lastly in Section 4 you will be asked 8 demographic questions.

#### Survey Instructions

As genetics providers who discuss genetic testing and obtain informed consent directly with patients, we are asking you to respond to a list of <u>minimum and critical components</u> for a patient to provide informed consent for the genetic test.

In this section you will be asked to read a set of scenarios and answer corresponding questions. The scenarios provided are intended to demonstrate a ‘targeted discussion’ approach to obtaining informed consent, and only contain the critical concepts that were identified in the previous Delphi consensus modeling study. In Section 3 you will be shown the complete list of those concepts and will be asked to further comment on the adequacy of the list.

Before you begin, we want to provide some context.

<u>Informed consent</u> is defined as the process of obtaining a patient’s authorization before conducting a healthcare intervention, in this case, genetic testing.

Each scenario provides an example of a ‘targeted discussion’ approach to obtaining informed consent.

- In these scenarios, please focus on CLINICAL genetic testing (not research consents).
- The content of these discussions are meant to be agnostic to <u>WHO</u> performs the discussion (e.g. a genetics provider or a non-genetics provider with appropriate content expertise and scope of practice), or <u>HOW</u> and <u>WHERE</u> they occur (this could happen face to face, over phone or via telehealth).
- This is a time-limited (10-20 minute appointment) conversation.
- Targeted discussions are not meant to replace “traditional genetic counseling” when indicated for complicated conditions or those that involve a potential risk for adverse psychological reactions (for example predictive testing for adult onset neurodegenerative conditions); those with a near term risk for mortality or significant morbidity in the diagnosis; or for a patient with significant anxiety or decisional conflict about the testing or who requests traditional genetic counseling.

A few additional thoughts about our process:

- Clinicians can always identify patients who would benefit from a more intensive communication process, either providing it themselves or making a referral.
- Patients should always be able to ask questions and clarifications, request more information or obtain traditional genetic counseling if desired.
- The list of topics you will be asked to comment on should be considered specific to the concepts that a “typical patient” would consistently need to know in order to make an informed decision about whether or not to undergo genetic testing.
- While traditional genetic counseling has often included information on the logistics of testing and its cost/payment, and we recognize that for some patients these are impactful parts of the decision making process, we are separating these important practical/logistical components from the critical components of informed consent.
- To focus on the informed consent process, the CADRe workgroup is also separating out a detailed discussion of the prognosis and management of a condition, especially for individuals that already have a clinical diagnosis, from the discussion of the genetic testing decision making.

#### Survey Scenarios - participants completed a random three out of six

Please read the following case scenarios and answer the following questions after each, keeping in mind the above information.

Scenario 1, LQTS:

A 30-year-old male has had a clinical diagnosis of Long QT syndrome (LQTS) for several years. He has a prolonged QT interval for which he is being managed medically and has a maternal family history of syncope including several relatives with implantable cardioverter-defibrillators (ICDs). He wants to pursue genetic testing for LQTS. His provider explains the following concepts as part of the pre-testing informed consent process:

- Genetic testing is optional.
- We are doing this test to look for a genetic cause of your LQTS. The genetic test evaluates the genes associated with LQTS.
- The test may be positive, meaning that we found a variant in a gene that is responsible for the LQTS.
- While you are already being treated for LQTS, a positive test could refine your medical management if a gene specific treatment becomes known. In some cases, a specific gene diagnosis can also change what we know about your prognosis.
- The results may impact your family in different ways (their health, emotions, or relationships), and you may want to share the results. For example, positive test results could be used for your family members to find the cause of their symptoms, to predict risk for relatives who do not have symptoms, and for family planning.
- The test has limits. Even though you have a clinical diagnosis of LQTS, the test may be negative, meaning that we may not find a genetic cause of your LQTS in the genes tested. Lastly it may be inconclusive, meaning that a variant was found but we do not know if it causes your LQTS or is normal genetic variation. We would discuss any possible next steps after results are available.
- The results would be reported directly to me and I will contact you with them by [manner and approximate timeline].
- While genetic information is protected against discrimination of employment or health insurance by federal law, it is not protected against discrimination for long term care or disability, or life insurance. This is most relevant for relatives without symptoms who choose to have genetic testing.

Scenario 2, Lynch syndrome:

A 60-year-old female is newly diagnosed with endometrial cancer and has a family history of colorectal, uterine, and gastric cancers. She does not meet the clinical diagnostic criteria for Lynch syndrome, but would like to pursue genetic testing for hereditary cancer. Her provider explains the following concepts as part of the pre-testing informed consent process:

- Genetic testing is optional.
- We are doing this test to look for a genetic cause for your personal and family history of cancer, which is suggestive of Lynch syndrome. The genetic test evaluates the genes associated with Lynch syndrome. It is possible to cast a wider net and test for additional genes associated with other types of hereditary cancer. But the current test will only focus on genes related to cancer risks.
- The test may be positive, meaning that we found a variant in a gene that is responsible for your cancer.
- While you are already being treated for endometrial cancer, a positive test for Lynch syndrome would come with screening recommendations for the other associated cancers, such as more frequent colonoscopies, and/or surgical recommendations. In some cases, a specific gene diagnosis can also change what we know about your prognosis.
- The results may impact your family in different ways (their health, emotions, or relationships), and you may want to share the results. For example, positive test results could be used for your family members to find the cause of their cancers, to predict risk for relatives who do not have symptoms, and for family planning.
- The test has limits. It may be negative, meaning that we did not find a genetic cause of your cancer in the genes tested. Lastly it may be inconclusive, meaning that a variant was found but we do not know if it causes your cancer or is normal genetic variation. In either case we would discuss any possible next steps after results are available.
- The results would be reported directly to me and I will contact you with them by [manner and approximate timeline].
- While genetic information is protected against discrimination of employment or health insurance by federal law, it is not protected against discrimination for long term care or disability, or life insurance. This is most relevant for relatives without symptoms who choose to have genetic testing.

Scenario 3, HBOC:

A healthy 25-year-old female has a family history of breast, ovarian, and pancreatic cancers for which no one has had genetic testing. She would like to evaluate her risk for developing cancer. Her provider explains the following concepts as part of the pre-testing informed consent process:

- Genetic testing is optional.
- We are doing this test to look for a genetic predisposition to cancer based on your family history of cancer. The genetic test evaluates the genes associated with hereditary breast and ovarian cancers. It is possible to cast a wider net and test for additional genes associated with hereditary cancers. But the current test will only focus on genes related to cancer risks.
- The test may be positive, meaning that we found a variant in a gene that puts you at greater risk for developing breast, ovarian, and potentially other cancers, and likely explains your family history of cancer.
- A positive test result for hereditary breast and ovarian cancer would come with screening recommendations depending on the gene identified, such as earlier and more frequent mammograms. For some genes, a surgery to remove the ovaries and reduce ovarian cancer risk may also be recommended after age 35.
- The results may impact your family in different ways (their health, emotions, or relationships), and you may want to share the results. For example, positive results on the test could be used for your family members to find the cause of their cancers, to predict risk for relatives who do not have symptoms, and for family planning.
- The test has limits. We may not find a genetic change associated with cancer risk in the genes tested, a ‘negative’ result’. It would still be possible that there is a genetic predisposition to cancer in your family members that you did not inherit, and they could consider testing. It would also be possible that there is a genetic cause for the cancers in your family that we can’t test for, and you would still be at an increased risk for cancer. If we do not find a genetic change in you, we will still recommend cancer screening based on your family history. Lastly it may be inconclusive, meaning that a variant was found but we do not know if it increases your risk for cancer or is normal genetic variation. In either case we would discuss any possible next steps after results are available.
- The results would be reported directly to me and I will contact you with them by [manner and approximate timeline].
- While genetic information is protected against discrimination of employment or health insurance by federal law, it is not protected against discrimination for long term care or disability, or life insurance. This is most relevant for people without symptoms who choose to have genetic testing.

Scenario 4, OTC deficiency:

A healthy 32-year-old female has a family history of ornithine transcarbamylase deficiency (OTC deficiency). Her sister recently had a son who was diagnosed with OTC deficiency. Genetic testing revealed a pathogenic variant in the X-linked *OTC* gene, inherited from his mother (your patient’s sister). The patient is planning to become pregnant and would like to be tested for the familial variant, since she understands she is at risk of having the variant herself. Her provider explains the following concepts as part of the pre-testing informed consent process:

- Genetic testing is optional.
- We are doing this test to determine if you inherited the familial *OTC* variant. The test only looks for the familial variant in the *OTC* gene. No other genes (or variants, depending on the test) will be tested.
- The test may be positive, meaning that you inherited the same OTC variant as your sister, although your risk for symptoms may be different than that of your family members. It may be negative, meaning that neither you or your children are at risk for OTC symptoms.
- If you are positive, there would be additional management recommendations for your own health, such as special care during and after pregnancy. In addition, the results of the test could help you understand what your chances are to have a child with OTC, and we could discuss the options available to you before and during a pregnancy if you are found to be positive.
- The results would be reported directly to me and I will contact you with them by [manner and approximate timeline].
- While genetic information is protected against discrimination of employment or health insurance by federal law, it is not protected against discrimination for long term care or disability, or life insurance. This is most relevant for relatives without symptoms who choose to have genetic testing.

Scenario 5, RASopathy:

A 1-year-old female is suspected to have a RASopathy. Her provider recommends a RASopathy gene panel to establish a diagnosis and the parents express interest. The provider explains the following concepts as a part of the pre-testing informed consent process:

- Genetic testing is optional.
- We are doing this test to look for a genetic explanation for her symptoms. The genetic test evaluates the genes associated with RASopathies, which cause a group of disorders that involve symptoms including facial features, heart defects, and developmental delay, like those seen in your daughter.
- The test may be positive, meaning that we found a variant in a gene that is responsible for her symptoms and provides a diagnosis of a specific disease.
- While she is already being treated by a cardiologist for her heart problem, a positive test and a specific diagnosis could help clarify whether any additional screenings or treatments would be helpful, such as an eye exam or speech therapy. In some cases, a specific gene diagnosis can also change what we know about your prognosis.
- The results may impact your family in different ways (their health, emotions, or relationships), and you may want to share the results. For example, positive results on the test could be used for your family members for relatives with similar symptoms, and for family planning.
- The test has limits. Even though it is suspected that she has a RASopathy, the test may be negative, meaning that we may not find a genetic cause of her features in the genes tested. Lastly it may be inconclusive, meaning that a variant was found but we do not know if it causes her features or is normal genetic variation. In either case we would discuss any possible next steps after results are available.
- The results would be reported directly to me and I will contact you with them by [manner and approximate timeline].
- While genetic information is protected against discrimination of employment or health insurance by federal law, it is not protected against discrimination for long term care or disability, or life insurance. This is most relevant for relatives without symptoms who choose to have genetic testing.

Scenario 6, Exome sequencing:

A 10-year-old male is evaluated for intellectual disability and dysmorphic facial features. His family history is negative. Previous negative testing includes fragile X and chromosomal microarray. His parents are motivated to pursue further testing to explain his developmental history. The provider recommends exome sequencing and explains the following concepts as a part of the pre-testing informed consent process:

- Genetic testing is optional.
- We are doing this test to look for a genetic explanation of his features. The genetic test evaluates all of the patient’s genes to look for a cause of his intellectual disability and dysmorphic facial features.
- The test may be positive, meaning that we found a variant in a gene that is responsible for his features. We would also learn how the variant is inherited, such as from one parent, both, or neither.
- While he is already receiving special education services, a positive test could help us determine if any other specialty services that may be useful, including evaluations for health concerns associated with the genetic cause. In some cases, a specific genetic diagnosis can help us know what to expect based on what we know about other people with the same genetic diagnosis.
- The results may impact your family in different ways (their health, emotions, or relationships), and you may want to share the results. For example, positive results on the test could be used for your family members for family planning. You may want to share the results.
- The test has limits. The test may be negative, meaning that we did not find a genetic cause of his features. The test may be inconclusive, meaning that a variant was found but we do not know if it caused his features or if it is normal genetic variation, though including parent samples in the analysis helps interpret these inconclusive results. If parent samples are included, you may learn unexpected information about family relationships. In either case we would discuss any possible next steps after results are available.
- Since this test evaluates all of the genes, it is an option to receive testing for other genes that cause other health problems, such as cancer and heart problems, for which there are established medical screenings or treatments available. It is your choice whether to include these genes in the test or not.
- The results would be reported directly to me and I will contact you with them by [manner and approximate timeline].
- While genetic information is protected against discrimination of employment or health insurance by federal law, it is not protected against discrimination for long term care or disability, or life insurance. This is most relevant for relatives without symptoms who choose to have genetic testing.

## REFERENCES

1. Attard CA, Carmany EP, Trepanier AM. Genetic counselor workflow study: The times are they a-changin‘? J Genet Couns. 2019;28(1):130–140.

2. Joseph G, Pasick RJ, Schillinger D, Luce J, Guerra C, Cheng JKY. Erratum to: Information Mismatch: Cancer Risk Counseling with Diverse Underserved Patients (J Genet Counsel, 10.1007/s10897-017-0089-4). J Genet Couns. 2017;26(5):1105.

3. Hitchcock EC, Study C, Elliott AM. Shortened consent forms for genome-wide sequencing: Parent and provider perspectives. Mol Genet Genomic Med. 2020;8(7):e1254.

4. Hallquist MLG, Tricou EP, Ormond KE, et al. Application of a framework to guide genetic testing communication across clinical indications. Genome Med. 2021;13(1):71.

5. Robson ME, Bradbury AR, Arun B, et al. American Society of Clinical Oncology Policy Statement Update: Genetic and Genomic Testing for Cancer Susceptibility. J Clin Oncol. 2015;33(31):3660–3667.

6. Committee Opinion No. 693: Counseling About Genetic Testing and Communication of Genetic Test Results. Obstet Gynecol. 2017;129(4):e96–e101.

7. Ormond KE, Borensztein MJ, Hallquist MLG, et al. Defining the Critical Components of Informed Consent for Genetic Testing. Journal of Personalized Medicine. 2021;11(12):1304.

8. Qualtrics software, Version Feb-March 2020 of Qualtrics. Copyright © 2020 Qualtrics. Qualtrics, Provo, UT, USA.

9. Ormond KE, Hallquist MLG, Buchanan AH, et al. Developing a conceptual, reproducible, rubric-based approach to consent and result disclosure for genetic testing by clinicians with minimal genetics background. Genet Med. 2019;21(3):727–735.

10. IBM Corp. Released 2019. IBM SPSS Statistics for Windows, Version 26.0. Armonk, NY: IBM Corp

11. Seaver LH, Khushf G, King NMP, et al. Points to consider to avoid unfair discrimination and the misuse of genetic information: A statement of the American College of Medical Genetics and Genomics (ACMG). Genet Med. doi:10.1016/j.gim.2021.11.002

12. Unim B, Pitini E, Lagerberg T, Adamo G, De Vito C, Marzuillo C, Villari P. Current Genetic Service Delivery Models for the Provision of Genetic Testing in Europe: A Systematic Review of the Literature. Front Genet. 2019 Jun 19;10:552. doi: 10.3389/fgene.2019.00552. PMID: 31275354; PMCID: PMC6593087.

